# Bidirectional relationship between olfaction and Parkinson’s disease

**DOI:** 10.1101/2023.10.18.23297218

**Authors:** Jonggeol J. Kim, Sara Bandres-Ciga, Karl Heilbron, 23andMe Research Team, Cornelis Blauwendraat, Alastair J. Noyce

## Abstract

**Background:** Hyposmia (loss of smell) is a common early symptom of Parkinson’s disease (PD). The shared genetic architecture between hyposmia and PD is unknown.

**Methods:** We leveraged genome-wide association study (GWAS) results for self-assessment of ‘ability to smell’ and PD diagnosis. Linkage disequilibrium score regression (LDSC) and Local Analysis of [co]Variant Association (LAVA) were used to identify genome-wide and local genetic correlations. Mendelian randomization was used to identify potential causal relationships.

**Results:** LDSC found that sense of smell negatively correlated at a genome-wide level with PD. LAVA found negative correlations in four genetic loci near *GBA1, ANAPC4, SNCA*, and *MAPT*. Using Mendelian randomization we found evidence for strong causal relationship between PD and liability towards poorer sense of smell, but weaker evidence for the reverse direction.

**Conclusions:** Hyposmia and PD share genetic liability in only a subset of the major PD risk genes. While there was definitive evidence that PD can lower the sense of smell, there was only suggestive evidence for the reverse. This work highlights the heritability of olfactory function and its relationship with PD heritability and provides further insight into the association between PD and hyposmia.

## Introduction

Parkinson’s disease (PD) is the second most prevalent neurodegenerative disease worldwide. The etiology of PD is complex; age, environmental and genetic risk factors, and comorbidities are all thought to contribute[1,2]. Genome-wide association studies (GWAS) have discovered multiple susceptibility loci for PD [3]. Impaired sense of smell (hyposmia) is a common early symptom of PD [4] but given that the onset of the disease precedes the diagnosis by years, if not decades, the direction of the causal relationship between hyposmia and PD is unclear. Although it is assumed that early deterioration of smell is a reflection of selective vulnerability to the disease process, an alternative hypothesis is that exposures that trigger loss of smell may lead to PD [5,6].

Mendelian randomisation (MR), utilizing summary statistics from GWAS, can be used to explore the causal nature of an association between an observed intermediate phenotype (such as smell loss) and a disease outcome (such as PD). In the current study, we assessed shared genetic architecture between self-reported sense of smell and PD, and then evaluated the potential direction of effect between sense of smell and PD. While a GWAS of sensory perception of smell has been done before [7], this included a relatively small number of participants and resulted in a 11 genome-wide significant hits across African American and European populations. Here we used a large GWAS of self-reported sense of smell as a proxy for hyposmia to investigate genetic overlap with PD on a local and genome-wide scale.

## Methods

### GWAS summary statistics

We used two GWAS summary statistics datasets for PD [3] and self-reported “ability to smell” (Figure 1). We used summary statistics from only clinically-ascertained datasets for PD which resulted in 15,056 cases and 12,637 controls. The contributing PD datasets have previously been described [3] but in brief: clinically ascertained unrelated PD cases without known Mendelian forms of PD were analyzed against neurologically healthy controls across 13 different studies using logistic regression. The results were then meta-analyzed using fixed-effect meta-analysis. Any variants with a heterogeneity statistic I^2^ greater than 80 and minor allele frequency less than 0.001 were removed.

**Figure 1.**
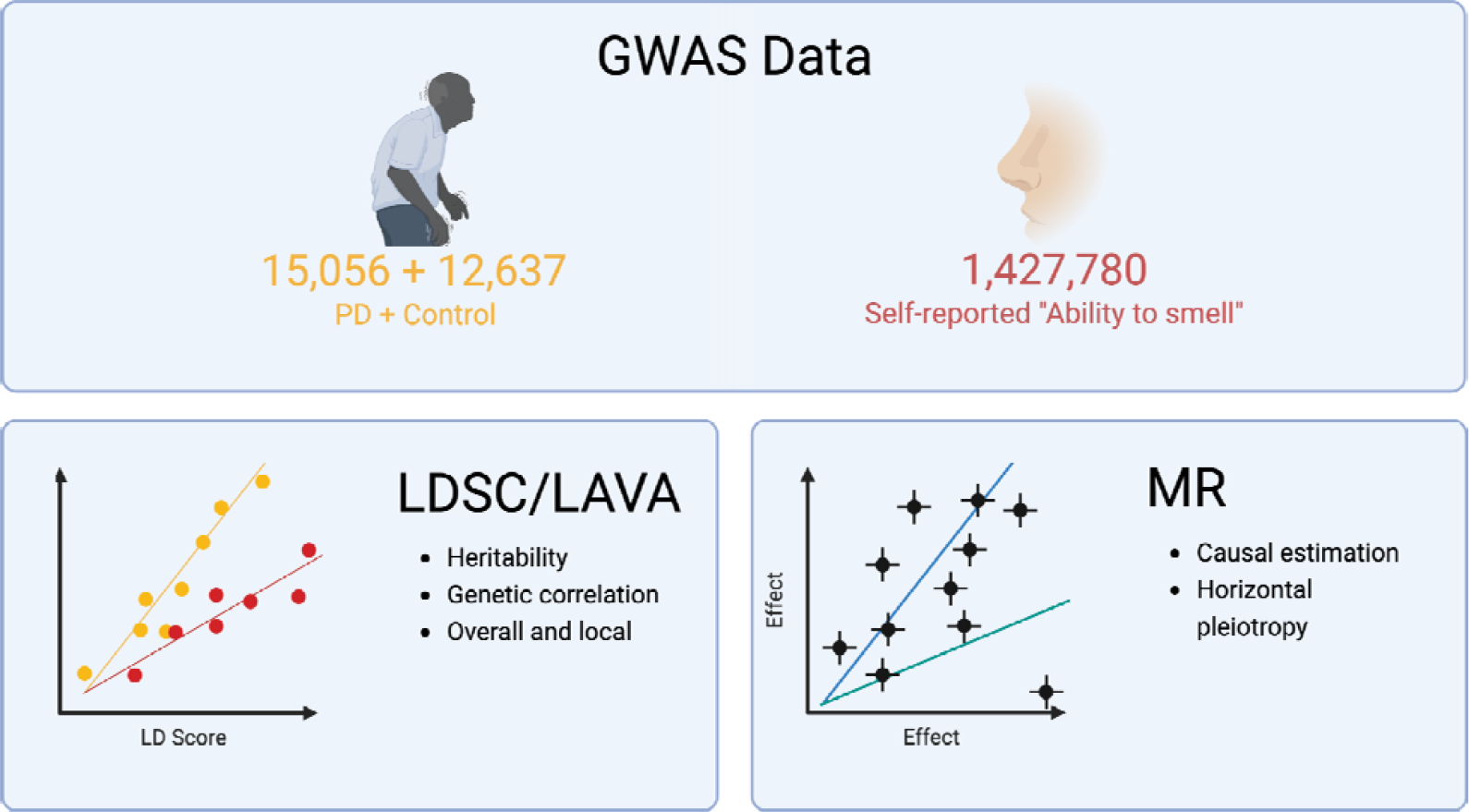
Study Design.

**Figure 2.**
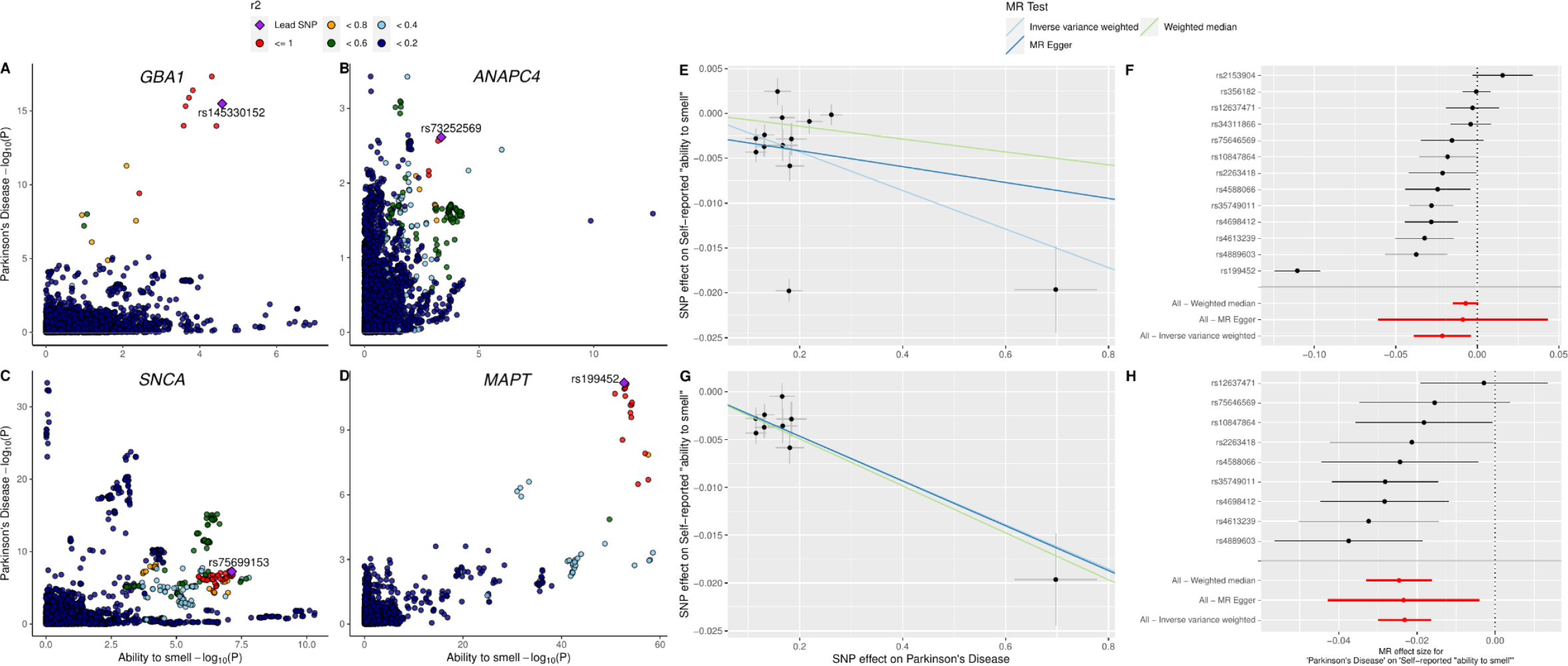
LocusCompare plots of LAVA results and MR results on PD and ability to smell. A-D) LocusCompare plots of *GBA1, ANAPC4, SNCA*, and *MAPT* loci with PD and ability to smell. Lead SNP is the SNP with the lowest sum of the P values from both studies. LD r^2^ value relative to the lead SNP is indicated by color. E-F) Scatter and forest plots of MR results before removal of the heterogeneity outliers. G-H) Scatter and forest plots of MR results after removal of heterogeneity outliers determined by MR-PRESSO.

### 23andMe, Inc. Ability to Smell GWAS

Detailed methods for the “ability to smell” GWAS can be found in the supplementary methods. In brief, self-reported data was collected online by 23andMe participants who provided informed consent. Participants were asked to rate their “ability to smell’’ on a 5-point scale from very poor (1) to very good (5). Participants provided informed consent and volunteered to participate in the research online, under a protocol approved by the external AAHRPP-accredited IRB, Ethical & Independent (E&I) Review Services. Related individuals were removed and to minimize confounding by ancestry, only individuals with predominantly European ancestry were used. Samples were genotyped, then were imputed using Beagle 5 [8] on an imputation panel that included Human Reference Consortium and custom 23andMe imputation panel. Association test results were computed by linear regression assuming additive allelic effects. Covariates for age, sex, and the top five genetic principal components (PCs) were included to account for residual population structure, and indicators for genotype platforms to account for genotype batch effects. The association test p-value reported was computed using a likelihood ratio test.

### Shared genetic architecture between hyposmia and PD

To determine the shared genetic heritability of PD and the ability to smell, we performed bivariate Linkage Disequilibrium Score (LDSC) regression [9]. The summary statistics for the two traits underwent standard quality control and harmonization. In brief, only common (minor allele frequency > 0.01) SNPs present in HapMap3 outside of the major histocompatibility complex (MHC) region were kept in the analysis. All variants with reference allele mismatch were removed.

Local Analysis of [co]Variant Association (LAVA) [10] was used to identify independent regions of the genome that showed high levels of correlation between the two traits. LAVA provides 2,495 independent genetic loci generated from European 1000 Genomes data [11] that minimizes linkage disequilibrium (LD) between each block. We analyzed 409 loci that had significant hits from our GWAS data and were able to be tested in both datasets without significant variant missingness. Bivariate LAVA was only run when both traits had significant univariate heritability in the locus, which was determined by a Bonferroni-corrected threshold (P_Heritability_ < 0.05/409). 55 loci were analyzed in the bivariate LAVA and significant results were determined using Bonferroni-corrected threshold of P < 0.05/55.

### Mendelian Randomization

Two-sample MR using the TwoSampleMR package in R (v4.1) [12,13] was used to determine the direction of effect between self-reported ability to smell and PD. Instrumental variables (IVs) for both phenotypes were selected by filtering for genome-wide significant variants (P < 5 x 10^-8^). The remaining variants were clumped (r^2^ < 0.001) to ensure independence. To generate and confirm strong instruments for the exposures, F-statistics were calculated for each IV. F-statistics were above 10, the recommended threshold for determining strong IVs[14]. We identified 13 and 276 variants as IVs for PD and smell respectively. The Wald ratios of the IVs were calculated and meta-analyzed using inverse variance weighted (IVW), MR-Egger, and weighted median (WM) methods. While IVW is the most powerful method for meta-analysis, MR-Egger is less biased in the presence of net directional pleiotropy and weighted median is a valid estimate when up to half the weight of the IVs are invalid. Heterogeneity of the Wald ratios was tested using Cochran’s Q statistic and converted to I^2^ which quantifies the level of heterogeneity. The Egger-intercept test was used to test for directional pleiotropy. To remove potential heterogeneity outlier IVs that may bias the results, MR-PRESSO [15] was used. The meta-analyses, Cochran’s Q, and Egger-intercept test were run again with the outliers removed and the results were compared with the previous results.

## Results

Here we describe the genetic correlation between self reported ability to smell and PD. The ability to smell was a highly polygenic trait with 394 genome-wide significant loci (Supplementary Table 1, Supplementary Figure 1). PD GWAS results contained 13 significant loci, a subset of the 90 loci discovered in the full GWAS which included self-reported and proxy cases [3]. The LDSC regression on the ability to smell GWAS results estimated a genomic control value of 2.1, but an LD score intercept of 1.056, indicating that the inflation was likely due to polygenicity rather than from population stratification, cryptic relatedness, or other biases. The observed scale heritability explained by common SNPs was 0.0541. We found that the single known risk variant for COVID-related anosmia was significant in the “ability to smell” GWAS data but the effect size was small (*UGT2A1/UGT2A2* locus rs7688383, Beta_Ability to smell_ = 0.0077, P = 1.2 x 10^-11^; OR_COVID-19 anosmia_ = 1.11, P = 1.4 x 10^-14^)[16].

**Table:**
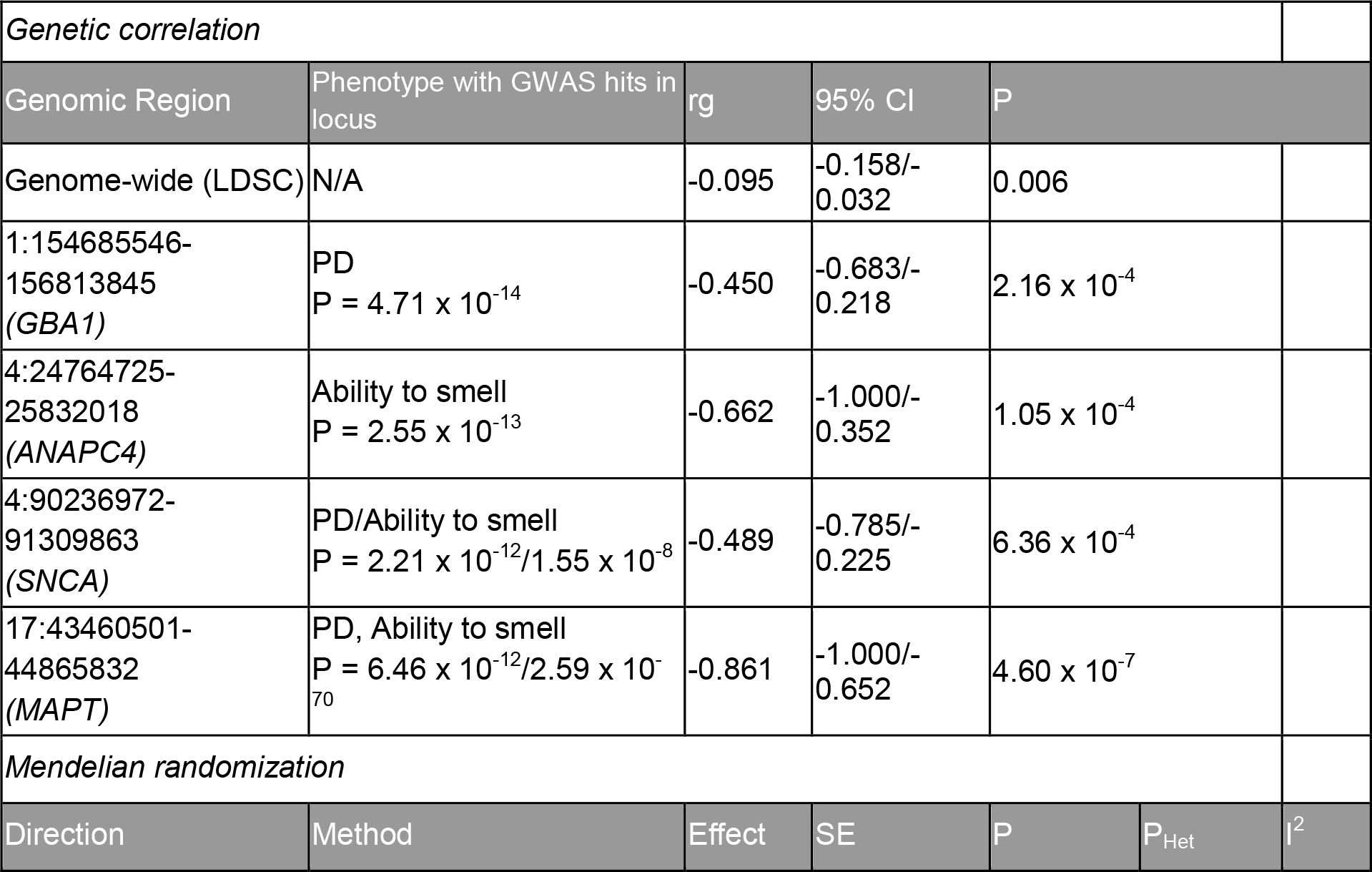
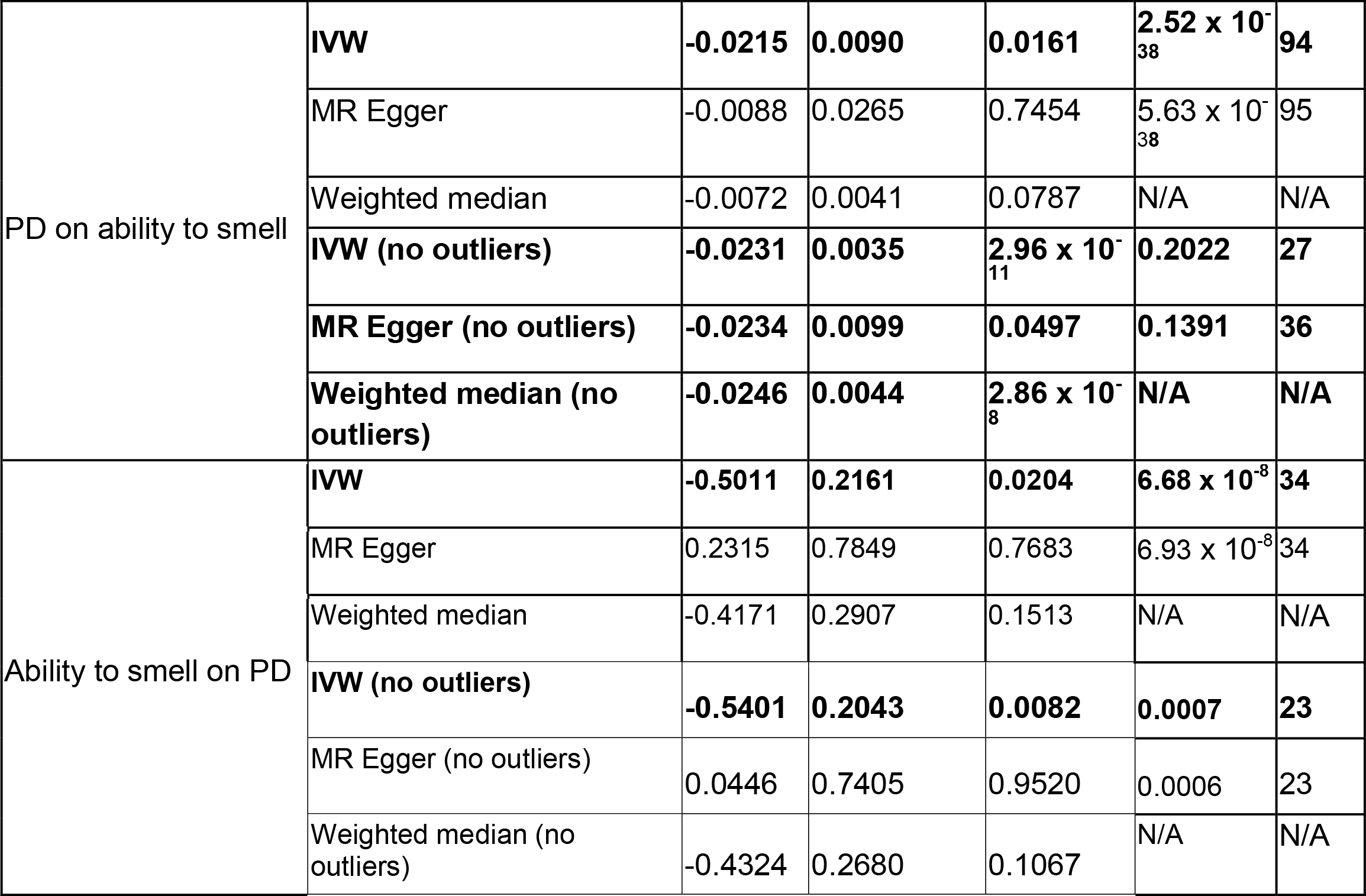
Genome-wide/local genetic correlation and MR results of PD on the self-reported ability to smell. Local correlation results that remained significant after Bonferroni correction are shown here. Genomic region is notated in *chromosome:basepair position start-basepair position end* in hg19 with relevant PD risk gene or the closest protein coding gene of the top GWAS hit. The P value in the second column represents the minimum P-value in the region for that phenotype. I^2^ and P_Het_ is N/A for Weighted median tests as the method does not test for heterogeneity. IVW - inverse variance weighted, PD - Parkinson’s disease. Significant MR results are **bolded**.

The genome-wide genetic correlation between the ability to smell and PD from LDSC revealed a significant inverse correlation (P = 0.006) such that hyposmia correlated with a higher risk of PD (Table). The correlation coefficient was modest at -0.095 (95% CI: -0.158/- 0.032). LAVA identified four local genetic correlations between PD and the ability to smell (Figure, Table). Two loci near *SNCA* and *MAPT* had significant GWAS hits in both traits, while two loci near *GBA1* and *ANAPC4* had significant hits in only PD and smell GWAS, respectively. All four loci showed negative correlation between the two traits. *MAPT* and *ANAPC4* loci contained -1.000 in its 95% confidence interval, suggesting potential collinearity.

Two-sample MR found evidence that liability towards PD was nominally associated with a reduction in self-reported ability to smell (Effect_IVW_ = -0.0215, P_IVW_ = 0.0161, Table, Figure). The instruments were highly heterogeneous (I^2^ > 80, Supplementary Table 2), but the Egger intercept test result was non-significant with the intercept close to 0 (intercept = -0.002, p = 0.6214 Supplementary Table 3). Although the *MAPT* variant rs199452 had significantly stronger effect size than others, a leave-one-out sensitivity analysis determined that the IVW result was not driven by any single variant (Supplementary Figure 2, Supplementary Table 4). MR-PRESSO was used to remove potential heterogeneity outliers which removed 4 out of 13 SNPs. These variants were near genes *SNCA, MAPT, TMEM175*, and *NUCKS1* (Supplementary Table 5). The remaining SNPs were near genes *GBA1, STK39, MCCC1, BST1, ELOVL7, LRRK2, HIP1R, SETD1A, RIT2*. Outlier removal increased the level of significance for all three meta-analysis methods and both IVW and WM were significant against multiple test corrected alpha threshold of 0.004 (P_IVW_ = 2.96 x 10^-11^, P_MR Egger_ = 0.0417, P_WM_ = 2.86 x 10^-8^). Heterogeneity between the instruments was no longer significant (P_Het_ > 0.05) and Egger intercept decreased further (intercept = -5 x 10^-5^, Supplementary Table 3). Leave-one-out analysis after MR-PRESSO also confirmed that the results were not driven by a single variant (Supplementary Figure 2).

The reverse direction MR result was also nominally significant (P_IVW_ = 0.0204, Supplementary Figure 3, Supplementary Figure 4) and not significant after multiple test correction (αBonferroni = 0.004). Leave-one-out test with IVW showed that the effect was not driven by a single variant (Supplementary Figure 2, Supplementary Table 4). The effect direction of IVW and MR-Egger results were different (Effect_IVW_ = -0.5011, Effect_MR Egger_ = 0.2315). While this often indicates possible bias due to net horizontal pleiotropy, the insignificant Egger intercept test (P = 0.332, Supplementary Table 3) and large confidence interval of the Egger effect size that overlaps with the IVW effect (95% CI_MR-Egger_: -1.307 to 1.770) suggests instead that the Egger estimation is imprecisely estimated and unable to test for horizontal pleiotropy. Both IVW and MR-Egger results showed a moderate level of heterogeneity (I^2^ > 30, Supplementary Table 2). Removing heterogeneity outliers increased the significance of the IVW results (P = 0.0082), but only borderline when corrected for multiple tests using Bonferroni correction (αBonferroni = 0.004). The MR-Egger effect direction continued to be different from IVW with a lower point estimate but large confidence interval (Effect_MR-Egger_ = 0.0446, 95% CI: -1.407 to 1.496, Supplementary Table 3). The Egger intercept values continued to be non-significant after MR-PRESSO (P = 0.570). While the heterogeneity was still present after MR-PRESSO (P_Het_ = 0.0007), the I^2^ indicated low levels of overall heterogeneity (I^2^ < 30).

## Discussion

Using genetic liability towards both traits, we first examined the genetic correlation between self-reported ability to smell and PD, and then used MR to examine the direction of the association. Olfaction is a complex system that involves multiple organs and tissues. The regions identified by the ability to smell GWAS might include variants that explain factors that influence the sense of smell in multiple ways, such as those that influence the olfactory bulb, nasal cavity, allergies, and propensity to smoke. Hyposmia caused by PD would represent a smaller subset of the potential pathways that contribute to olfaction, which may explain the relatively low global genetic correlation coefficient but strong local correlation in few select loci. PD-related hyposmia likely involves pathways related to degradation of olfactory bulbs or neurons present in the olfactory system. Previous studies have linked Lewy body pathology in the olfactory system with hyposmia [17], indicating direct degradation of the olfactory bulbs. Another possibility is that hyposmia is a by-product of neurodegeneration in the brain like other sensory deficits in PD. Indeed MRI imaging in PD patients have found neurodegeneration in both the olfactory bulb and brain regions associated with olfactory function [18,19]. The loci highlighted by LAVA further strengthen this theory, as *GBA1* variants are commonly associated with worse cognitive outcomes in PD patients [20,21] and it has been suggested that *MAPT* is associated with dementia in PD [22]. Worse cognitive clinical presentation due to these variants is likely to coincide with hyposmia. PD patients with *LRRK2* mutations typically have milder cognitive decline [23], which may explain the lack of correlation in the *LRRK2* region.

*ANAPC4* locus, one of the regions highlighted by LAVA, has not been previously identified as a potential risk factor for PD. The lead SNP for this locus in the ability to smell GWAS is rs34811474 (P < 2.55 x 10^-13^), which is a missense variant for *ANAPC4*, encoding the anaphase promoting complex subunit 4 protein. As the name suggests, the main function of the gene product is its involvement in anaphase in mitosis and meiosis. *ANAPC4* is relatively intolerant of loss-of-function and missense, with probability of being loss-of-function intolerant (pLI) score of 0.001 and missense constraint Z-score of 2.5[24]. However, given that rs34811474 is common in the non-Finnish European population (MAF = 0.2194)[24], the missense caused by this variant is likely benign. It is unclear how *ANAPC4* may influence the sense of smell or PD risk, but this variant is a lead SNP in other complex phenotypes such as body mass index [25] and urate levels [26], suggesting it is an influential and complex region.

The MR analysis suggested a causal association between PD and hyposmia. Interestingly MR-PRESSO removed variants near *SNCA, MAPT, TMEM175*, and *NUCKS1* as heterogeneity outliers, independently identifying *SNCA* and *MAPT* regions as potential pleiotropic regions. As these two regions were also identified as having high genetic correlation by LAVA, these regions may cause hyposmia more directly through an unknown pathway via horizontal pleiotropy, while other regions with high PD risk such as *LRRK2* contribute to hyposmia indirectly by contributing to PD risk. These variants need to be further investigated for a shared mechanism of action which may have an impact on diagnosis and prodromal disease progression.

When using MR in the reverse direction, we did not find strong evidence to support the possibility of a bidirectional effect, where a poorer sense of smell is also a potential risk for PD. The two IVW tests were nominally significant and MR-PRESSO IVW was borderline after Bonferroni-correction. Given the large sample size discrepancy between the two traits (15,056 PD cases vs 1.4 million 23andMe participants), the borderline results suggest a future PD GWAS with a larger clinically-ascertained sample may be needed to find a conclusive result. Nevertheless, factors that cause loss of smell have been previously hypothesized to cause neurodegenerative diseases. One potential vector is exposure to air pollutants which have been linked to brain inflammation and neurodegenerative disease pathology [27,28]. Another is the infiltration of a neurotropic pathogen via the olfactory system, which is a crucial part of the Braak hypothesis for PD [29,30]. Influenza [5] and hospitalization due COVID-19 infection [31] have previously been identified as potential risk factors for PD. A recent study comparing α-synuclein seed amplification assays (SAA) of nasal brushings and skin biopsies found that the distribution of α-synuclein deposition is not uniform across PD patients [32]. The study supported a hypothesis based on two subtypes of PD: “body-first,” where pathogenic α-synuclein starts aggregation in the enteric nervous system and spreads to the brain, and “brain-first,” where it begins in the brain and then affects the rest of the body [33]. The heterogeneity and lower significance of the reverse MR results could be attributed to such clinical heterogeneity.

There are several limitations to this study. First, studies have shown that self-assessment of hyposmia is biased when compared to clinical measurements [34] which may introduce self-report bias to our study. In addition COVID-19 infection cases were not omitted from the “ability to smell” data however this is likely to be a small subset of the population included in the 23andMe data. We found that the single known risk variant for COVID-related hyposmia (*UGT2A1/UGT2A2* locus rs7688383) was significant in sense of smell data (P = 1.2 x 10^-11^) but had a very small effect size (Beta = 0.0077). Lastly, this study only looked at data in European populations. There is limited evidence that different ancestries present varying clinical manifestations of PD [35]. While there are currently efforts to diversify PD genetics, there is a dearth of data in more underrepresented populations. Global Parkinson’s Genetics Program (GP2) is an initiative that seeks to sequence over 150,000 PD participants globally [36]. We will need in depth research of the colocalized regions in diverse populations to better understand the genetic relationship between hyposmia and PD.

## Data Availability

The full GWAS summary statistics for the 23andMe discovery data set will be made available through 23andMe to qualified researchers under an agreement with 23andMe that protects the privacy of the 23andMe participants. Datasets will be made available at no cost for academic use. Please visit https://research.23andme.com/collaborate/#dataset-access/ for more information and to apply to access the data.

## Acknowledgements

A.J.N. reports grants from Parkinson’s UK, Barts Charity, Cure Parkinson’s, NIHR, Innovate UK, Virginia Keiley benefaction, Alchemab, Aligning Science Across Parkinson’s Global Parkinson’s Genetics Program (ASAP-GP2) and Michael J Fox Foundation.

We thank all members of the International Parkinson Disease Genomics Consortium (IPDGC). For a complete overview of members, acknowledgements, and funding, please see the Supplemental data and/or http://pdgenetics.org/partners.

We thank the research participants and employees of 23andMe. The following members of the 23andMe Research Team contributed to this study:

The following members of the 23andMe Research Team contributed to this study: Stella Aslibekyan, Adam Auton, Elizabeth Babalola, Robert K. Bell, Jessica Bielenberg, Jonathan Bowes, Katarzyna Bryc, Ninad S. Chaudhary, Daniella Coker, Sayantan Das, Emily DelloRusso, Sarah L. Elson, Nicholas Eriksson, Teresa Filshtein, Pierre Fontanillas, Will Freyman, Zach Fuller, Chris German, Julie M. Granka, Karl Heilbron, Alejandro Hernandez, Barry Hicks, David A. Hinds, Ethan M. Jewett, Yunxuan Jiang, Katelyn Kukar, Alan Kwong, Yanyu Liang, Keng-Han Lin, Bianca A. Llamas, Matthew H. McIntyre, Steven J. Micheletti, Meghan E. Moreno, Priyanka Nandakumar, Dominique T. Nguyen, Jared O’Connell, Aaron A. Petrakovitz, G. David Poznik, Alexandra Reynoso, Shubham Saini, Morgan Schumacher, Leah Selcer, Anjali J. Shastri, Janie F. Shelton, Jingchunzi Shi, Suyash Shringarpure, Qiaojuan Jane Su, Susana A. Tat, Vinh Tran, Joyce Y. Tung, Xin Wang, Wei Wang, Catherine H. Weldon, Peter Wilton, Corinna D. Wong.

K.H. is a former employee of 23andMe Inc., and holds stock and stock options in 23andMe, Inc.

This work was supported by the following grants and institutions: Intramural Research Program of the NIH, National Institute on Aging (NIA) (project numbers ZIAAG000534 and 1ZIAAG000534 to S.B.C, C.B.), National Institutes of Health, Department of Health and Human Services (C.B.); National Institute of Neurological Disorders and Stroke (project numbers ZO1 AG000535 and ZIA AG000949 to C.B.) This research was funded by Aligning Science Across Parkinson’s through the Michael J. Fox Foundation for Parkinson’s Research (MJFF).

Figure 1 was created using Biorender.com

This research utilised Queen Mary’s Apocrita HPC facility, supported by QMUL Research-IT. http://doi.org/10.5281/zenodo.438045

## Code and Data Availability

All code related to this manuscript will be available on the GP2 Github repository and deposited on Zenodo.

