## Supplementary Figures for "Bidirectional relationship between olfaction and Parkinson’s disease"

### Supplementary Figure 1 - Manhattan Plots of a) PD and b) ability to smell GWAS

a.

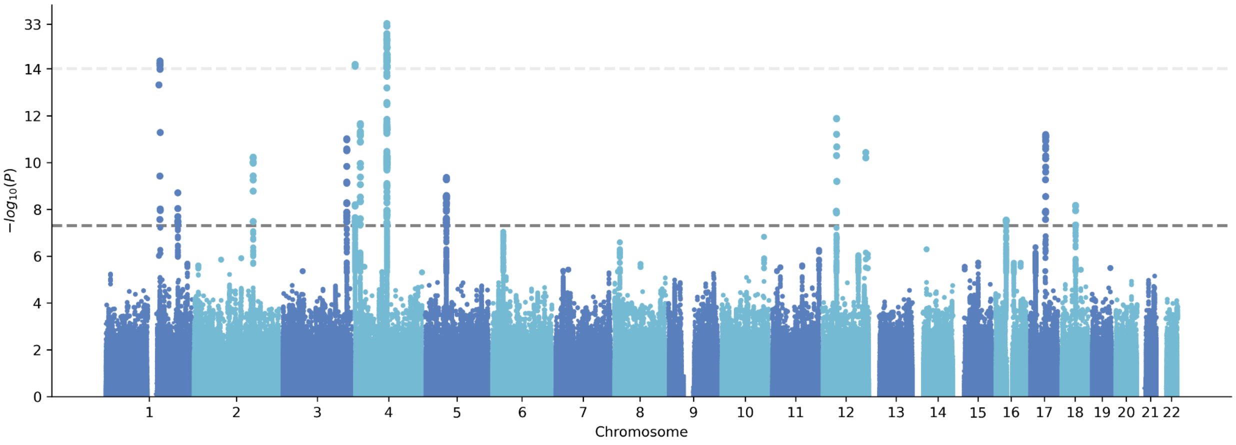


b.

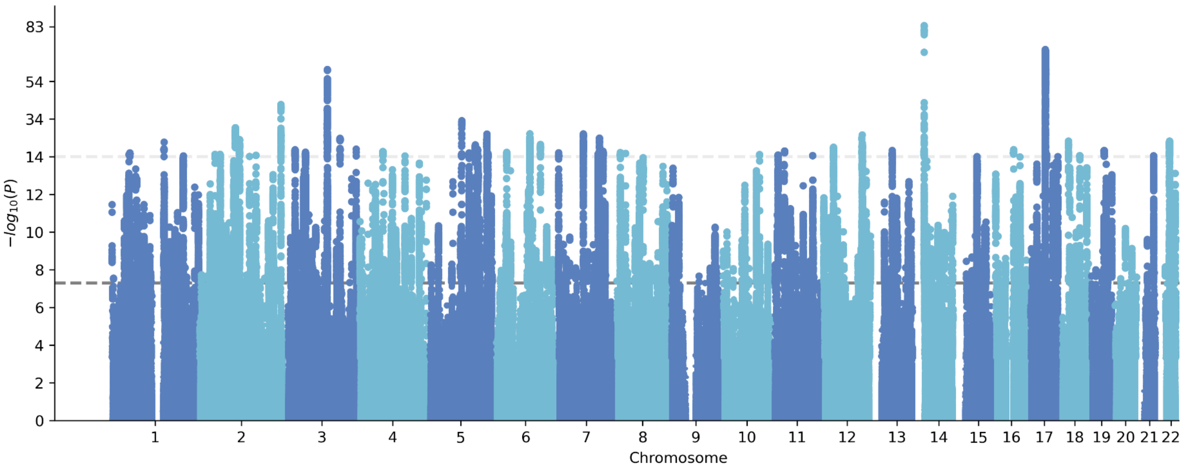

### Supplementary Figure 2 - Forest plot of leave-one-out sensitivity results

#### PD on ability to smell

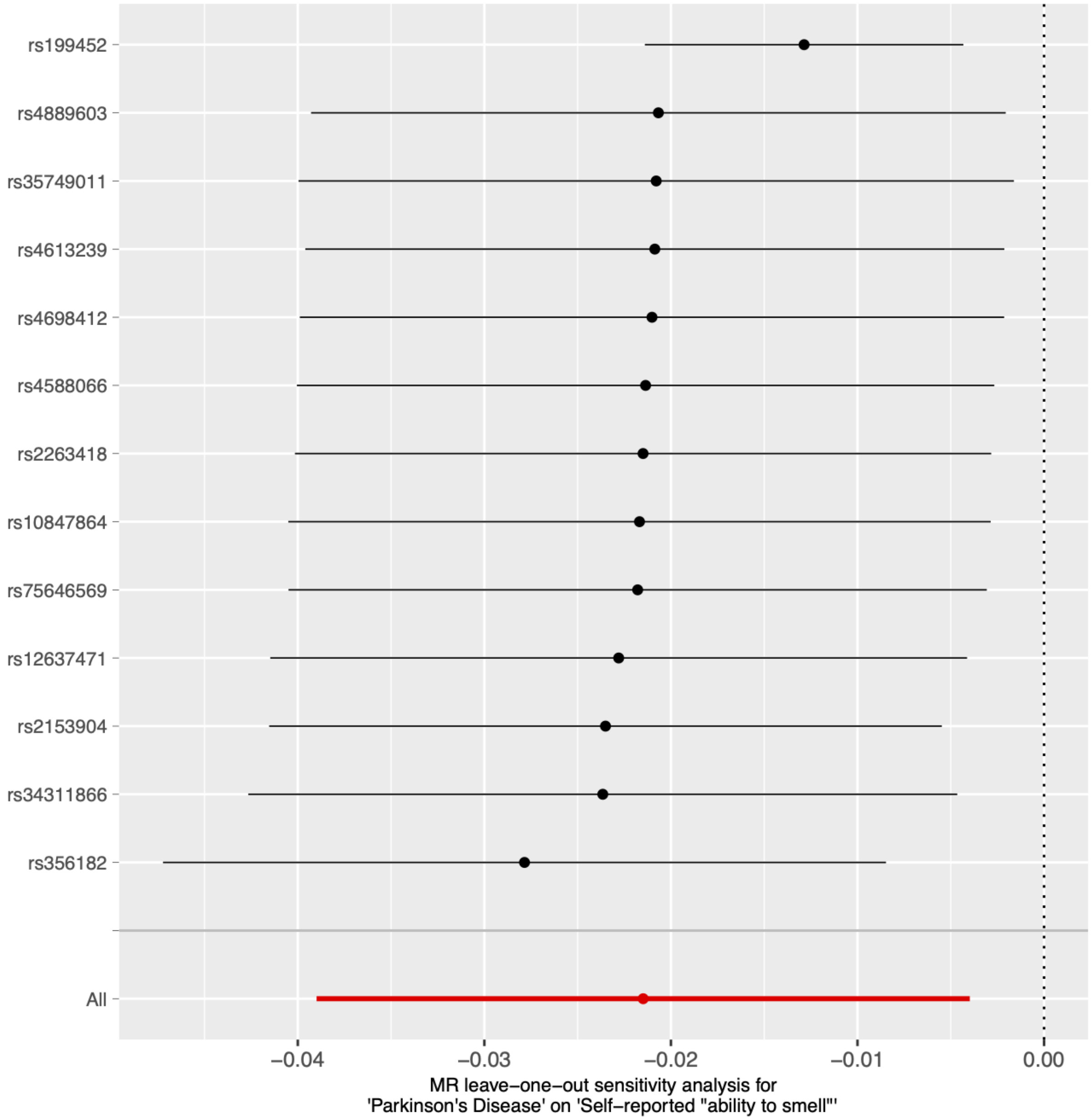

PD on ability to smell (outliers removed)

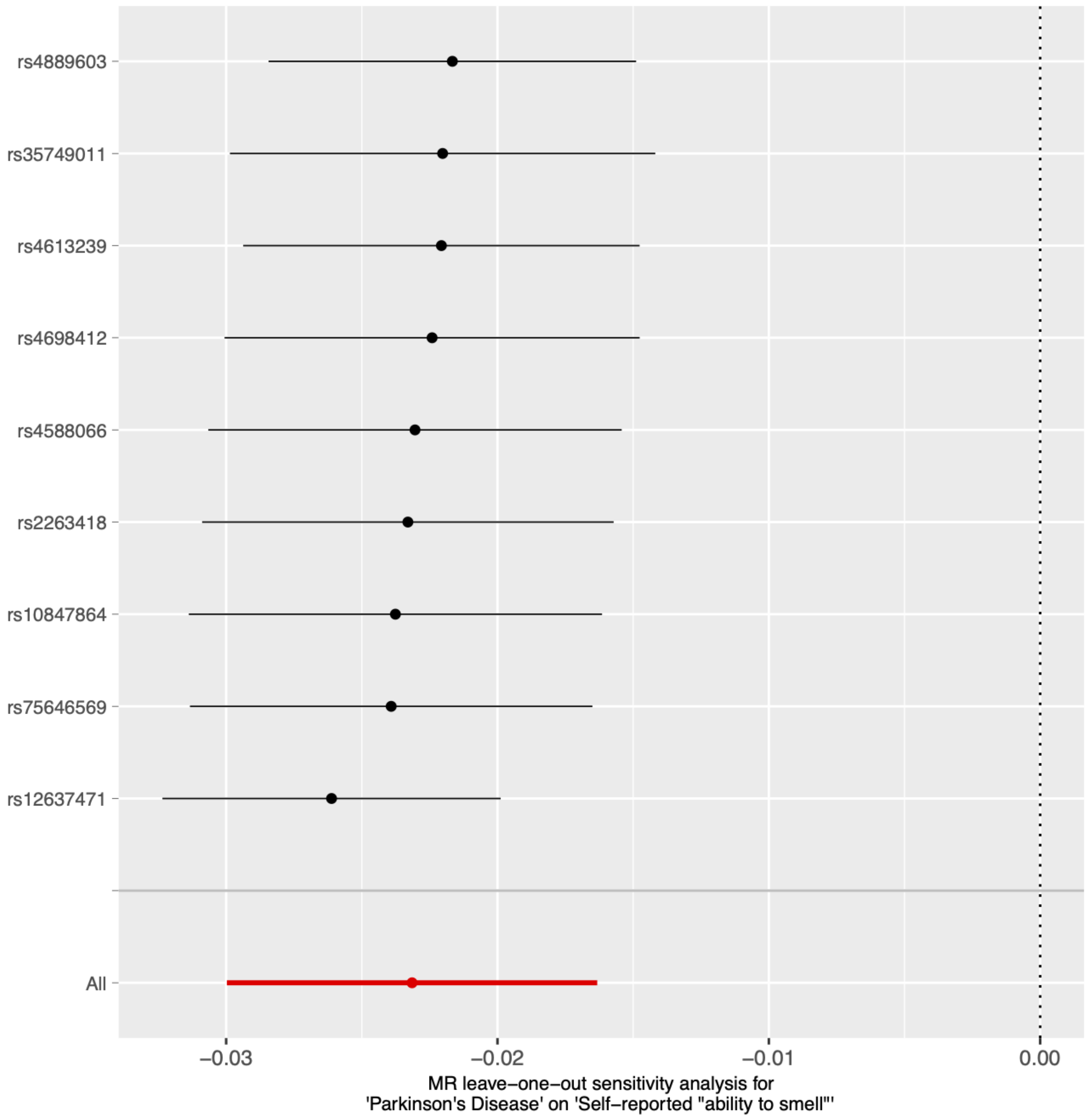

Ability to smell on PD

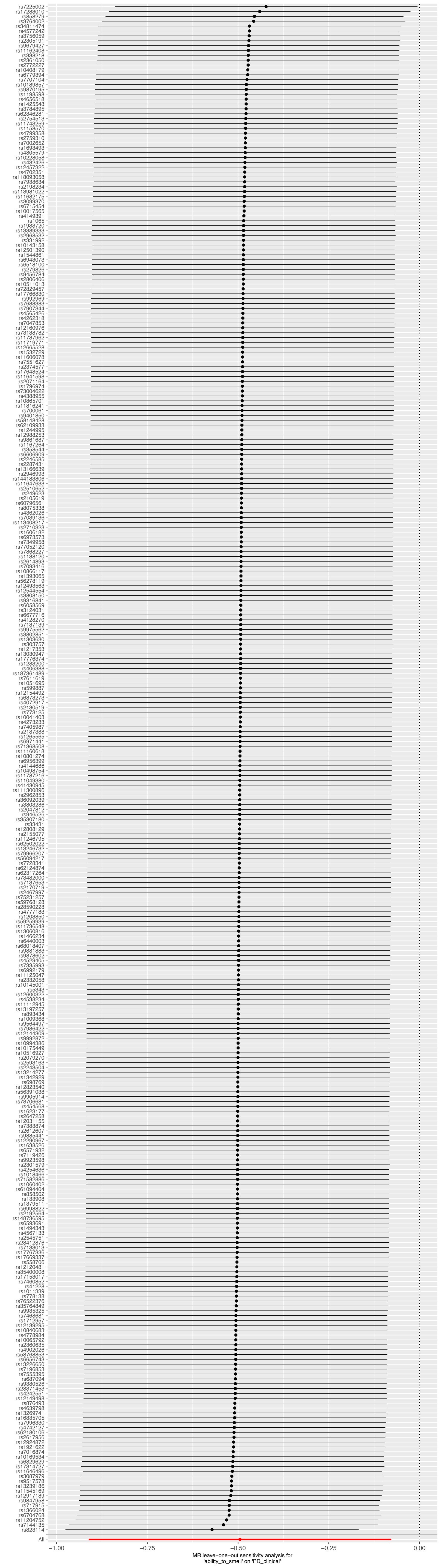

All
−1.00
−0.75
−0.50
−0.25
0.00
MR leave−one−out sensitivity analysis for
'ability_to_smell' on 'PD_clinical'

Ability to smell on PD (outliers removed)

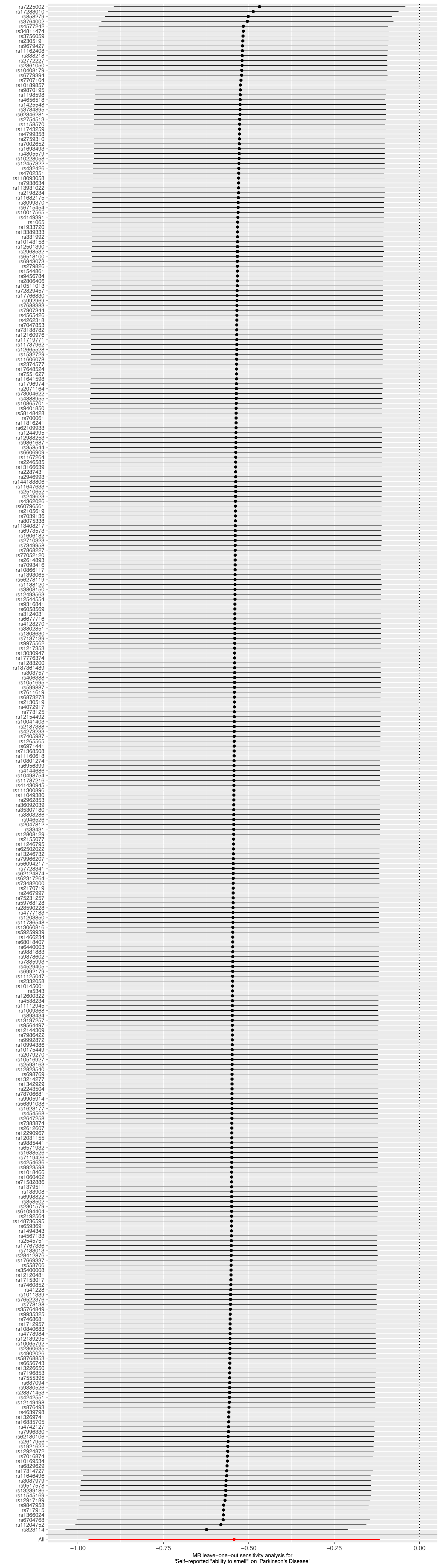

All
−1.00
−0.75
−0.50
−0.25
0.00
MR leave−one−out sensitivity analysis for
'Self−reported "ability to smell"' on 'Parkinson's Disease'

Supplementary Figure 3 - Forest plot of reverse-causation MR results

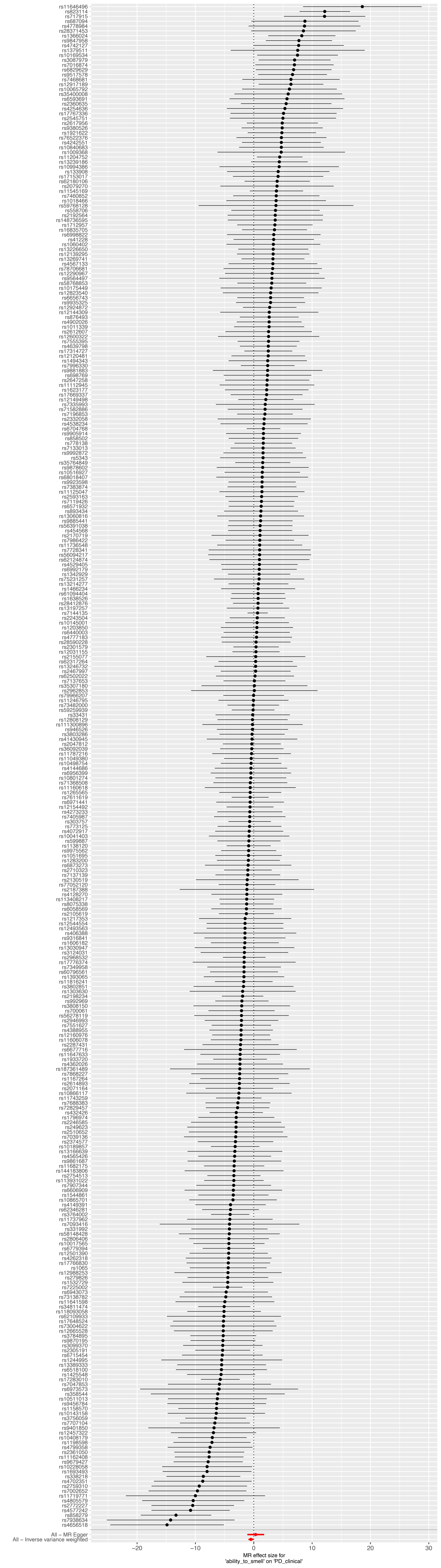

All – MR Egger
All – Inverse variance weighted
−20
−10
0
10
20
30
MR effect size for
'ability_to_smell' on 'PD_clinical'

Supplementary Figure 4 - Scatter plot of reverse-causation MR results

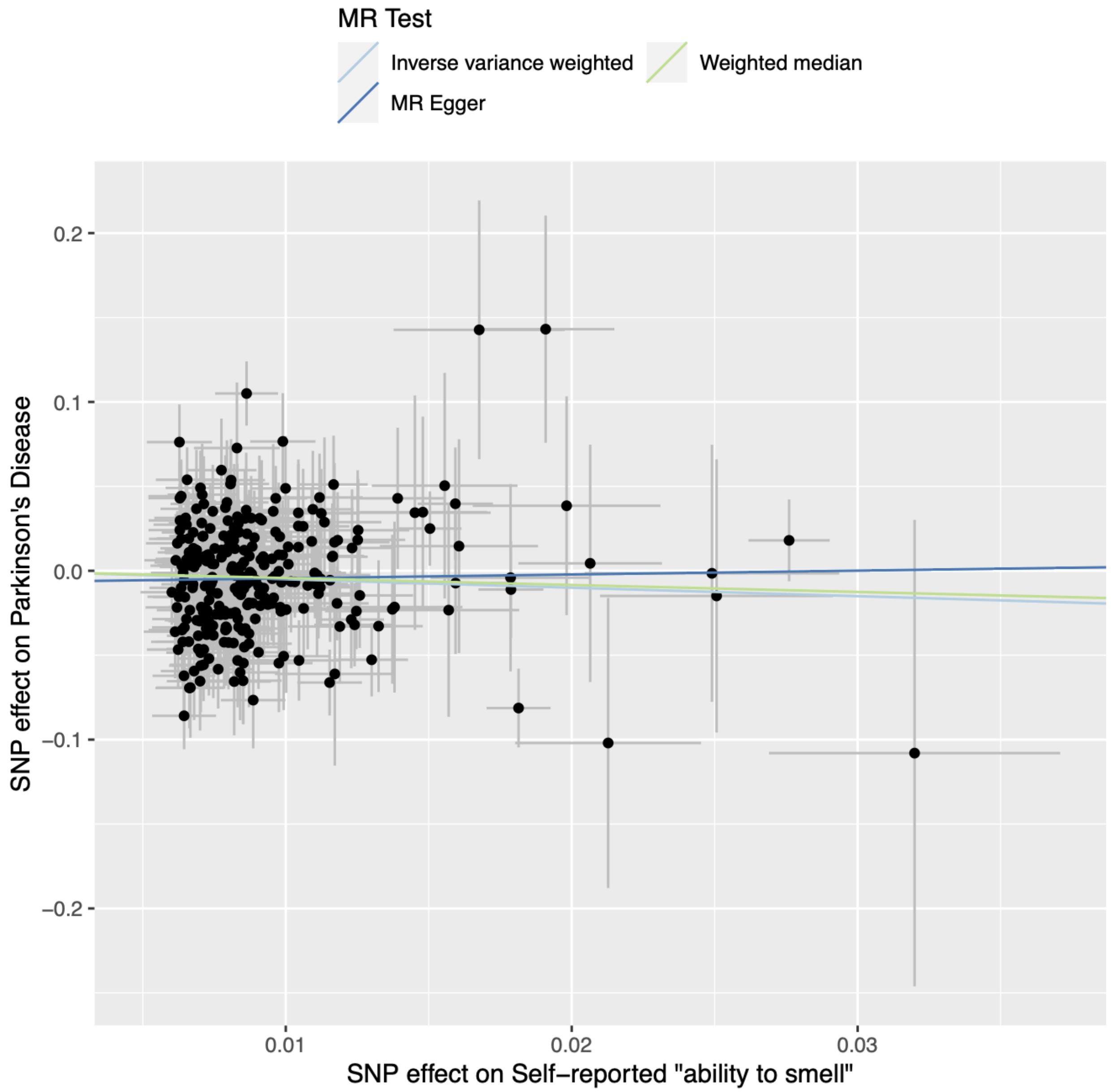

Supplementary Figure 5 - Scatter plot of outlier-removed reverse-causation MR results

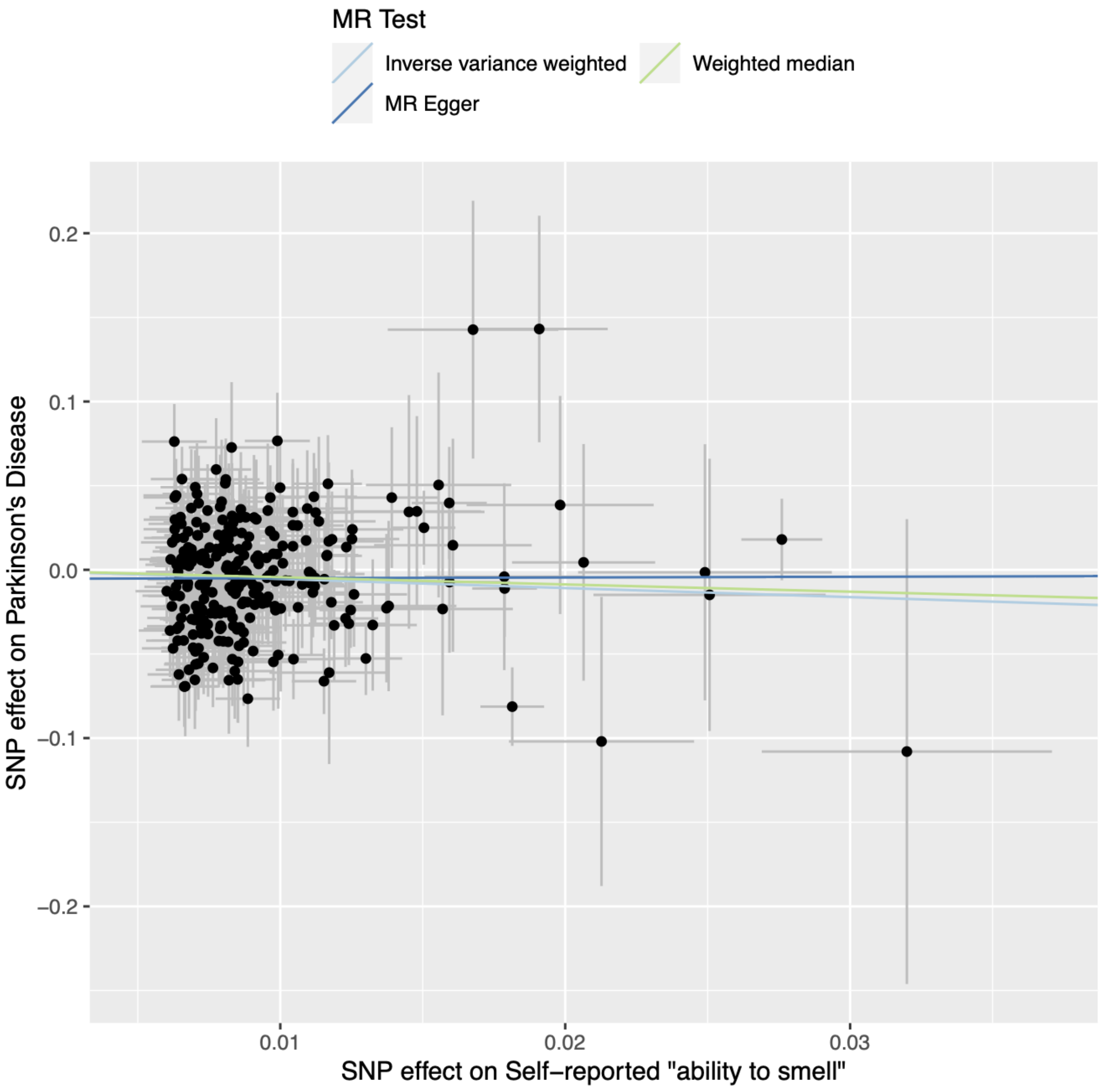
